# MedRAGent: An Automatic Literature Retrieval and Screening System Utilizing Large Language Models with Retrieval-Augmented Generation

**DOI:** 10.1101/2025.09.18.25335860

**Authors:** Zhuoyi Chen, Tianyi Liu, Yangrui Mo, Qishen Fu, Sibin Lei, Tiejun Tong, Xiaoyu Tang

## Abstract

**Background:** Systematic reviews play a critical role in synthesizing evidence across numerous studies, providing a foundation for informed decision-making in medical practice. However, the process is resource-intensive, requiring proficiency in constructing Boolean queries and screening extensive literature, which are time-consuming and susceptible to inconsistencies, especially for non-expert researchers. While large language models (LLMs) offer a potential solution, their tendency to generate inaccurate or hallucinated content restricts their direct application in systematic reviews.

**Objective:** This study introduces and evaluates MedRAGent, a novel system that integrates LLMs with retrieval-augmented generation (RAG), designed to automate and enhance the efficiency and accuracy of Boolean query formulation and title/abstract screening in systematic reviews.

**Methods:** MedRAGent employs DeepSeek-V3-0324 and Kimi-K2-0711-preview LLMs within an RAG framework tailored for PubMed. The system utilizes the official Medical Subject Headings (MeSH) database to construct precise Boolean queries. For screening, it employs the LLMs with a structured prompt to automatically evaluate the relevance of retrieved articles based on predefined inclusion and exclusion criteria. Its performance was assessed using 53,054 articles from 6 research topics.

**Results:** Our results showed that MedRAGent achieved an overall precision of 0.0271, recall of 0.8308, and F1-score of 0.0525 in Boolean query construction. For automated literature screening, the system attained an overall sensitivity of 0.8131, specificity of 0.9891, and G-mean of 0.8968 when using DeepSeek-V3-0324 as the underlying LLM. Performance improved when using Kimi-K2-0711-preview, with sensitivity of 0.8582, specificity of 0.9919, and G-mean of 0.9226. It efficiently processed 4,000-7,000 articles per day at low operational cost.

**Conclusions:** MedRAGent demonstrates strong potential for automating Boolean query construction and abstract-level screening in systematic reviews. It effectively accelerates literature processing, supporting researchers in conducting efficient and evidence-based medical reviews.

## Introduction

Systematic reviews and meta-analyses represent the highest standard of evidence-based research, forming the cornerstone of clinical guidance and policy decisions.^1^ However, the implementation of systematic reviews and meta-analyses faces numerous technical complexities. Conducting a comprehensive and systematic literature search requires proficiency in the retrieval rules of the database, the use of Boolean operators to construct the retrieval expression, the screening of the literature based on the title and abstract, and the exclusion of irrelevant studies. The process requires a significant investment of time and expertise, especially for novice researchers.

The precision of the literature search depends on the effective construction of Boolean queries, which are used to define specific search strategies to ensure that all relevant studies are covered while excluding irrelevant literature. However, due to the complex retrieval rules of the database and the large number of subject terms and free-text words related to the research questions, constructing Boolean queries manually is not only a very time-consuming and labor-intensive step,^2^ but also may not always meet researchers’ retrieval expectations. Wang et al. found that researchers adopted various methods to construct Boolean queries, and the reported recalls ranged from 41.38% to 91.28%.^3^ Current methodologies for constructing Boolean queries are predominantly manual and expert-driven, falling into two categories: the conceptual method^4^ and the objective method.^5^ In practice, researchers frequently combine the two techniques to compensate for their respective limitations, but this hybrid strategy further compounds the complexity and time required for query development.

In addition, the preliminary screening of literature based on titles and abstracts is crucial in the process of systematic review.^1^ Manual screening imposes substantial demands on time and human resources, with a single reviewer typically requiring approximately one hour to assess between 60 and 120 papers.^6^ Moreover, the screening process is prone to human errors, for example, omission of relevant literature and false inclusion of irrelevant literature.^7^ Due to fatigue and cognitive load, the rate of these errors may rise as the number of papers increases.^8^

Recent advances in large language models (LLMs) suggest possibilities for automating query design and screening. These models can interpret natural language instructions, propose search formulas, and rapidly evaluate text. However, LLMs are susceptible to producing inaccurate outputs—a phenomenon known as "hallucination"—when their training data contains errors, outdated information, or lacks domain-specific knowledge. Hallucination refers to the content generated by the model that appears plausible but is in fact incorrect, inconsistent with established facts, or logically flawed.^9^ For example, Wang et al. observed that ChatGPT generated Boolean queries containing invalid Medical Subject Headings (MeSH) terms. Even using optimized prompt, 55% of the MeSH terms generated by ChatGPT were not present in the official MeSH vocabulary, resulting in low recall and poor reproducibility.^3^

To mitigate LLM hallucinations and enhance performance, Gabin and Parapar utilized retrieval-augmented generation (RAG) to suggest alternative terms to users.^10^ These keyword-based suggestions significantly enhanced the efficiency and accuracy of document retrieval. RAG refers to the model’s process of retrieving relevant information from a large document corpus when answering questions or generating text. Subsequently, it uses the retrieved information to generate responses or texts, thereby improving the quality of predictions.^11^ Godinez proposed HySemRAG, a framework that combined the extraction, transformation, and loading pipeline with retrieval-augmented generation to synthesize large-scale documents and detected research gaps in specialized domains.^12^

Recent studies have investigated the application of LLMs for the automated retrieval and screening of scientific literature. Wang et al. designed five different prompt words for constructing Boolean queries using LLMs, and demonstrated that while ChatGPT could construct queries with high precision, their recall was significantly inferior to manually built queries.^3^ This shortcoming was confirmed by Staudinger et al. whose LLM-based pipeline using multiple open-source and closed-source LLMs to automatically create Boolean queries also produced queries with unacceptably low recall.^13^ Some studies applied LLMs to abstract screening. Guo et al. reported high aggregate sensitivity (0.76) and specificity (0.91) when screening over 24,000 clinical abstracts with GPT-4, but performance markedly declined at the individual dataset level.^14^ Similarly. Khraisha et al. used GPT-4 to screen abstracts and titles and observed consistently high specificity (>0.84) but sensitivity below 0.5 across diverse literature types.^15^ In contrast, a tailored approach by Matsui et al. achieved more stable and acceptable performance. By developing a specialized three-layer screening method, they utilized GPT-4 to effectively screen abstracts for systematic reviews on bipolar disorder, demonstrating both reliable sensitivity and specificity.^8^ This research highlights the potential of LLMs for clinical reviews, though further validation with larger and more diverse datasets is required.

In this paper, we propose MedRAGent, a novel literature retrieval and screening system that integrates LLMs with RAG to enhance the efficiency and accuracy of systematic reviews. The system employs LLMs for critical tasks including keyword extraction, synonym matching, and preliminary literature screening. To mitigate the challenge of LLM hallucination, MedRAGent incorporates a RAG framework that enables the system to retrieve relevant information from authoritative documents during text generation, substantially improving output reliability and accuracy.^11^ For example, MeSH terms are drawn directly from the controlled MeSH vocabulary, which ensures that the resulting Boolean queries are reproducible and semantically precise and offers greater control than free-text search alternatives. Tailored specifically for the PubMed database, MedRAGent operates through a structured workflow: the system first translates user-provided natural language prompts into formal Boolean queries, executes the literature retrieval, and then performs automated abstract-level relevance screening on the retrieved articles. We evaluated MedRAGent’s performance using data of 53,054 articles records from 6 research topics. We compared the system’s screening decisions against independent assessments by two human reviewers at the abstract level. We aim to determine MedRAGent’s effectiveness in constructing valid Boolean queries and to evaluate its capability to accurately identify relevant titles and abstracts in real-world clinical review datasets.

## Methods

MedRAGent, a based on web and local document retrieval-augmented generation system, is used for Boolean query construction and automated literature screening. The underlying LLMs use DeepSeek-V3-0324 and Kimi-K2-0711-preview, and the embedding model uses Pro/BAAI/bge-m3. The system consists of five stages. The first stage involves constructing the main index and supplementary index to source potential MeSH terms and Supplementary terms. The second stage involves retrieving and generating MeSH terms and Supplementary terms most relevant to the user’s query. The third stage expands the synonymous terms of MESH terms and Supplementary terms using the Medical Subject Headings Resource Description Framework (Mesh RDF) API. The fourth stage builds Boolean queries based on the user-defined construction rules, suffix rules, and retrieval time. The fifth stage conducts the literature retrieval and abstract-level relevance screening. In the following sections, we will provide in-depth explanations for each of these stages using the example of a systematic review on Ramadan fasting among adolescents with type 1 diabetes.^16^ Figure 1 shows the workflow of MedRAGent. The system is coded in Python.

**Figure 1.**
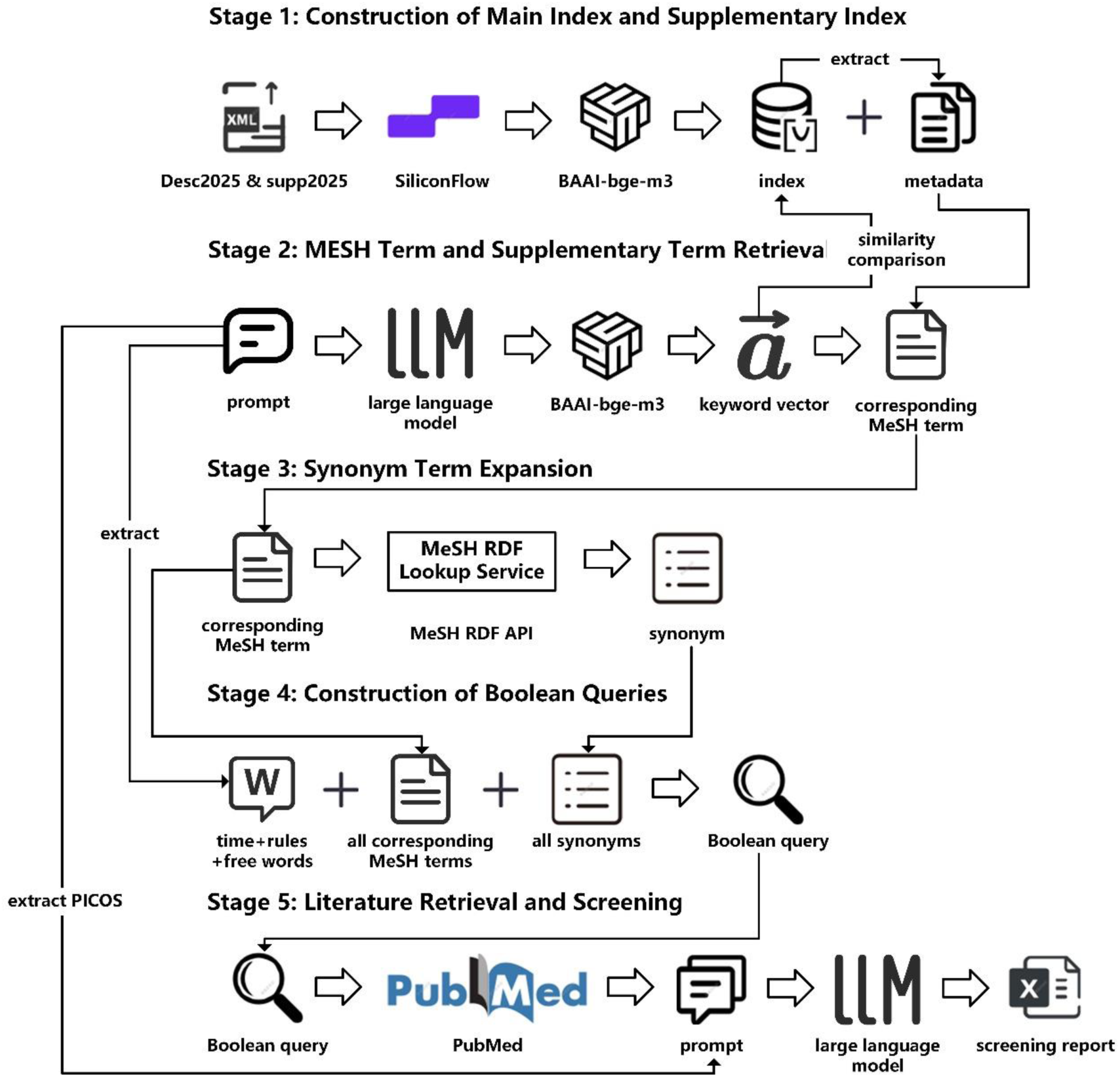
Workflow of MedRAGent.

### Stage 1: Construction of Main Index and Supplementary Index

The initial step involves constructing an index for the retrieval of relevant MeSH and supplementary terms. We download the files of Medical Subject Headings and Supplementary Concept Records (desc2025.xml and supp2025.xml) from the official website of the U.S. National Library of Medicine (NLM). MeSH is the controlled vocabulary used by NLM for indexing articles for PubMed. Supplementary Concept Records (SCRs) are the supplementary term lists of MeSH for new concepts (e.g., chemicals, drugs, proteins) that have not been incorporated into the annual MeSH review. We parse the XML files of MeSH and SCRs to extract the identifiers (UI), names, and classification numbers of each record and combine them into structured text. To ensure optimal performance in the Facebook AI Similarity Search (FAISS), vectors are L2-normalized (converted to unit vectors), making cosine similarity calculations dependent solely on vector direction rather than magnitude. Then, we add the normalized vectors to the FAISS to generate the main index and metadata for MeSH (mesh_index.faiss and mesh_metadata.pkl), and the supplementary index and metadata for SCRs (supple_index.faiss and supple_metadata.pkl), which are stored permanently on the disk.

### Stage 2: MESH Term and Supplementary Term Retrieval

First, we use LLM with a prompt to extract keywords from the given query and then use the embedding model to convert the keywords into query vectors, the prompt is shown in eTextbox 1 in Supplement 1. After L2 normalization, cosine similarity between two unit vectors û and v̂ simplifies to the dot product: 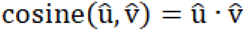. Candidate terms are ranked according to their cosine similarity scores relative to the query vector. A predefined similarity threshold of 0.6 determines the retrieval outcome. The search is first performed in the main MeSH index: if the highest-scoring vector meets or exceeds the threshold, the corresponding MeSH term is extracted from the metadata. If no match satisfies the threshold in the main index, the search proceeds to the supplementary index under the same condition. If the top candidate in the supplementary index also falls below the threshold, an empty value is returned. The eTextbox 2 in Supplement 1 shows the process for the example.

### Stage 3: Synonym Term Expansion

To improve the recall of our Boolean queries, we expand the initial set of MeSH and Supplementary Terms by gathering their synonyms. This is accomplished programmatically using the MeSH RDF API, a semantic data service provided by the NLM. For each term, we first retrieve its unique ID via the API and then use that ID to obtain a full list of its synonymous terms, including qualifiers and related descriptors. This process is illustrated in eTextbox 3 in Supplement 1.

### Stage 4: Construction of Boolean Queries

In this stage, the remaining keywords that do not match the specialized terms in the query are treated as free words and combined with the retrieved terms to form a Boolean query. The query is constructed using a defined set of rules for suffixes, Boolean operators, and a publication date filter. Suffix rules allow terms to be targeted to specific fields. For example, adding "[Title/Abstract]" to limit the search for the target keyword and term only in the article title and abstract fields. The Boolean construction follows either a default or custom rule. The default rule connects synonymous terms with “OR” and distinct concepts with “AND”; the custom rule allows the user to define these relationships manually. A publication date filter, based on a pre-specified time range, is then applied. This stage is detailed in eTextbox 4 in Supplement 1.

### Stage 5: Literature Retrieval and Screening

We conduct a literature search in PubMed using the constructed Boolean query through the Entrez Programming Utilities (E-utilities) API. To accommodate the API’s limitation of returning a maximum of 10,000 documents per request, we first determine the total number of results for each query. If this number exceed 10,000, we segment the retrieval process by year to avoid exceeding the limit; otherwise, documents are retrieved in a single operation. We then apply a LLM with a predefined prompt to perform relevance screening on the titles and available abstracts through API calls, the predefined prompt is shown in eTextbox 5 in Supplement 1. To mitigate the risk of excluding potentially relevant studies, we propose a "default inclusion" principle during literature screening. Under this principle, if no explicit evidence supporting inclusion or exclusion is found in the title or abstract, the record is automatically considered to meet the inclusion criteria and not meet exclusion criteria. This approach helps reduce false negatives that may arise from incomplete information.

### Data Analysis

We selected 6 published systematic reviews and meta-analyses, comprising a total of 53,054 literature records. These reviews covered a range of medical topics and varied in their screening scale, Boolean query strategies, and specific screening criteria. For our study, we collected their titles, abstracts, Boolean queries, and related evaluations. The population, intervention, comparison, outcome, study design and exclusion criteria of these 6 reviews are shown in eTable 1-6 in Supplement 1.

We first evaluated the Boolean query construction ability of MedRAGent. In this study, the number of retrieved records represents the total results returned by the MedRAGent-generated Boolean query, the number of relevant records is the number of articles identified as relevant during abstract screening by authors from the 6 published systematic reviews, and the number of relevant records retrieved is the subset of retrieved records that are relevant. These values are used to calculate precision, recall, and F1-score using the following formulas.^17^

Precision measures the proportion of relevant records among total results retrieved by MedRAGent and is calculated as:

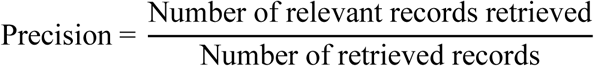

Recall measures the proportion of relevant records retrieved by MedRAGent and is calculated as:

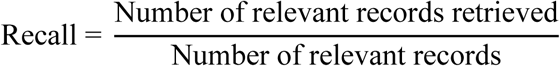

The F1-score, which provides a single metric balancing precision and recall, is the harmonic mean of the two:

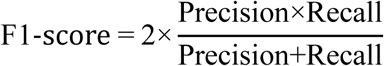

We then assessed MedRAGent’s abstract-level relevance filtering capability by comparing its inclusion/exclusion decisions directly against the gold standard dataset. To establish the gold standard dataset for assessment, two independent reviewers screened the titles and abstracts of all retrieved articles based on the predefined screening criteria and made inclusion/exclusion decisions for each entry. Any conflicts between reviewers were resolved through consensus negotiation. The performance was evaluated using sensitivity, specificity, and the G-mean. Sensitivity measures MedRAGent’s performance in correctly selecting records for inclusion and is calculated as:

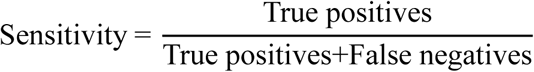

In our context, true positives are records that were included by both MedRAGent and reviewers, and false negatives are records that were included by reviewers but excluded by MedRAGent.

Specificity measures MedRAGent’s performance in correctly rejecting records for exclusion and is calculated as:

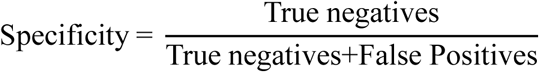

where true negatives are records that were excluded by both MedRAGent and reviewers, and false positives are records that were included by MedRAGent but excluded by reviewers.

The G-mean, which provides a single metric balancing sensitivity and specificity, is the geometric mean of the two:

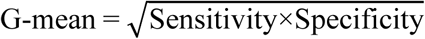

## Results

MedRAGent achieved an overall precision of 0.0271, recall of 0.8308, and an F1-score of 0.0525 in Boolean query construction (Table 1). Performance varied across individual studies. The query for Ramadan fasting among adolescents with type 1 diabetes yielded the highest precision (0.5227) while the query for vasodilators in acute heart failure resulted in the lowest precision (0.0081).^16,18^ Recall was generally robust across studies, with four out of six queries achieving recall values above 0.75. The highest recall was observed in the study on pharmacological interventions for insomnia disorder,^19^ which reached 0.9399.

**Table 1.**
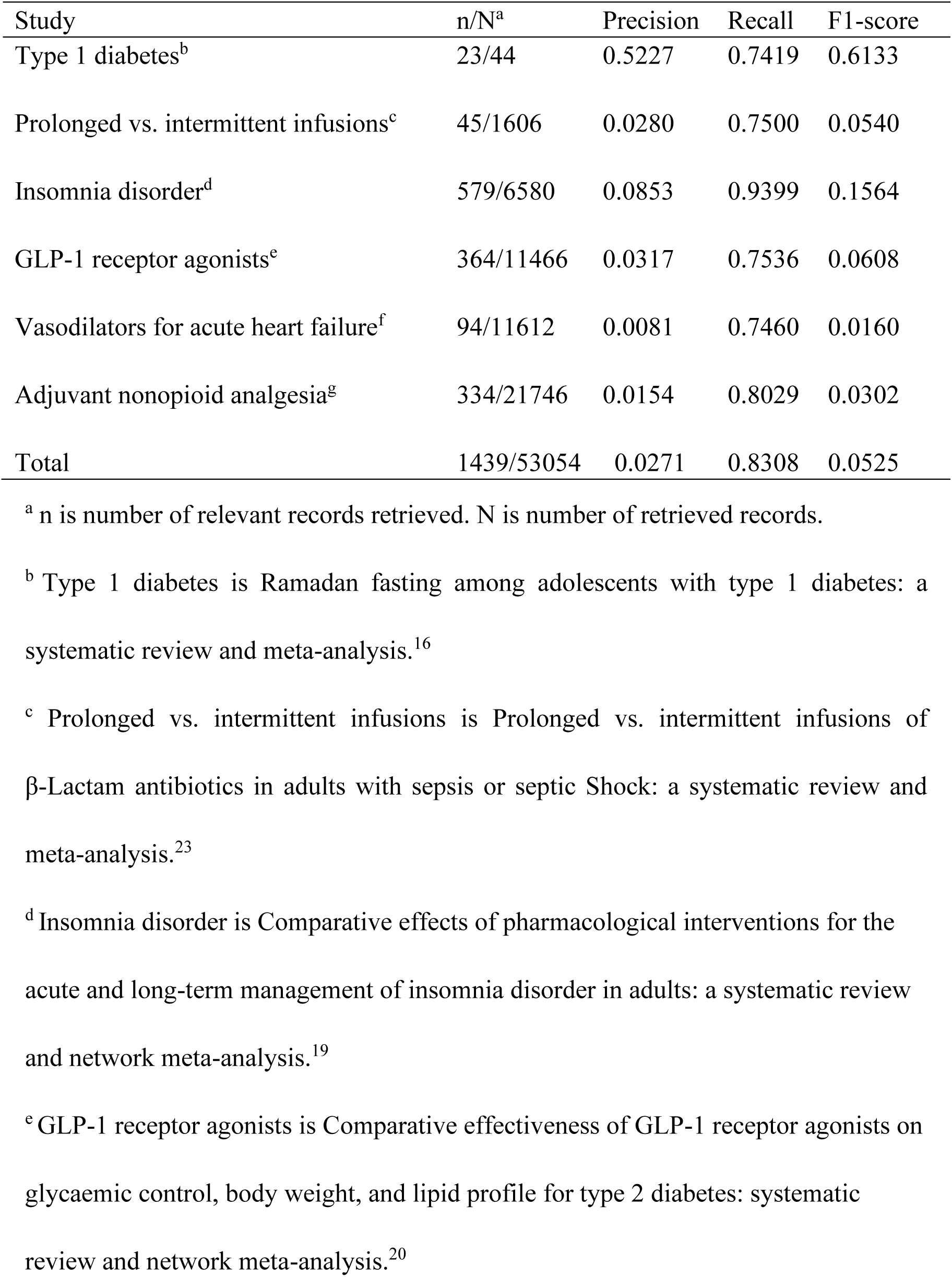
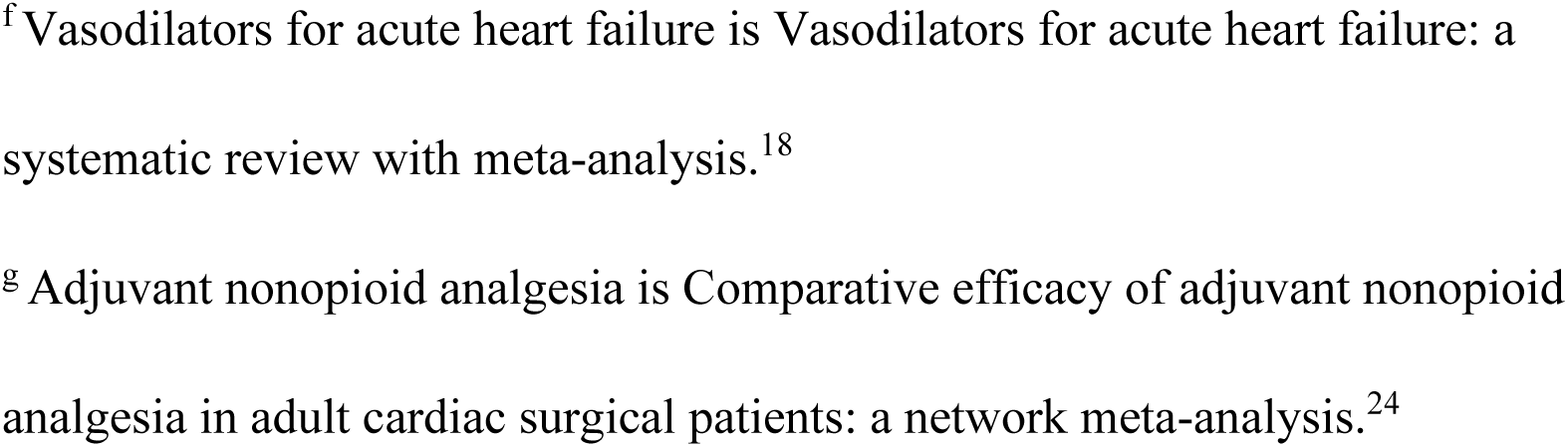
Performance of MedRAGent in Retrieving Articles.

In abstract-level screening, 50,599 articles with available abstracts were evaluated because a small portion of the literature abstracts were missing in the metadata exported by the E-utilities API. MedRAGent with DeepSeek-V3-0324 attained a sensitivity of 0.8131, specificity of 0.9891, and G-mean of 0.8968 (Table 2). Using Kimi-K2-0711-preview, it achieved higher performance, with sensitivity of 0.8582, specificity of 0.9919, and G-mean of 0.9226 (Table 3). The specificity values exceeded 0.97 in five studies. Ramadan fasting study showed slightly lower specificity (0.9286) compared to other topics.^16^ Sensitivity remained above 0.8 in most studies while exhibited reduced values (<0.8) by both models in the study on GLP-1 receptor agonists for type 2 diabetes.^20^

**Table 2.**
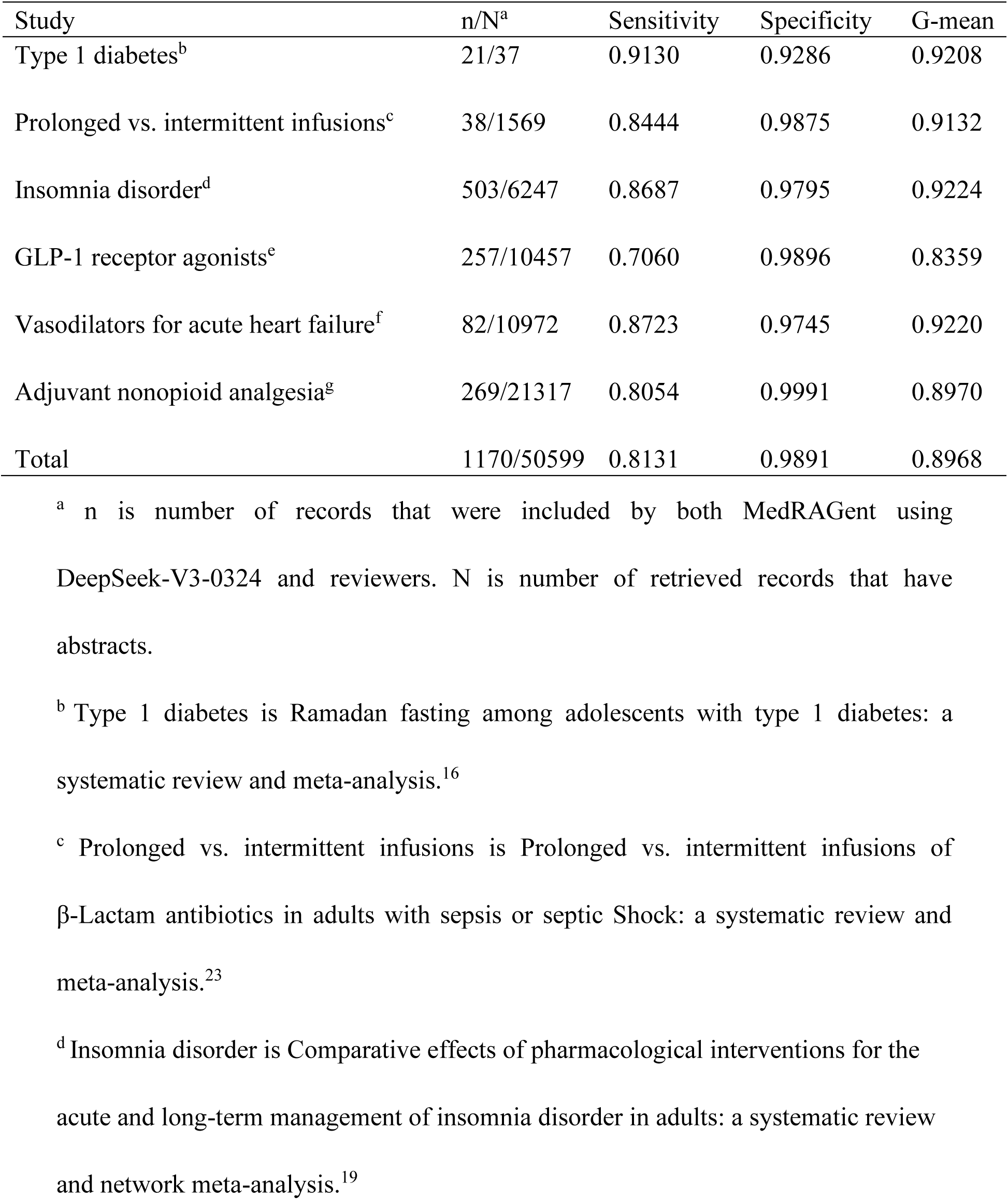
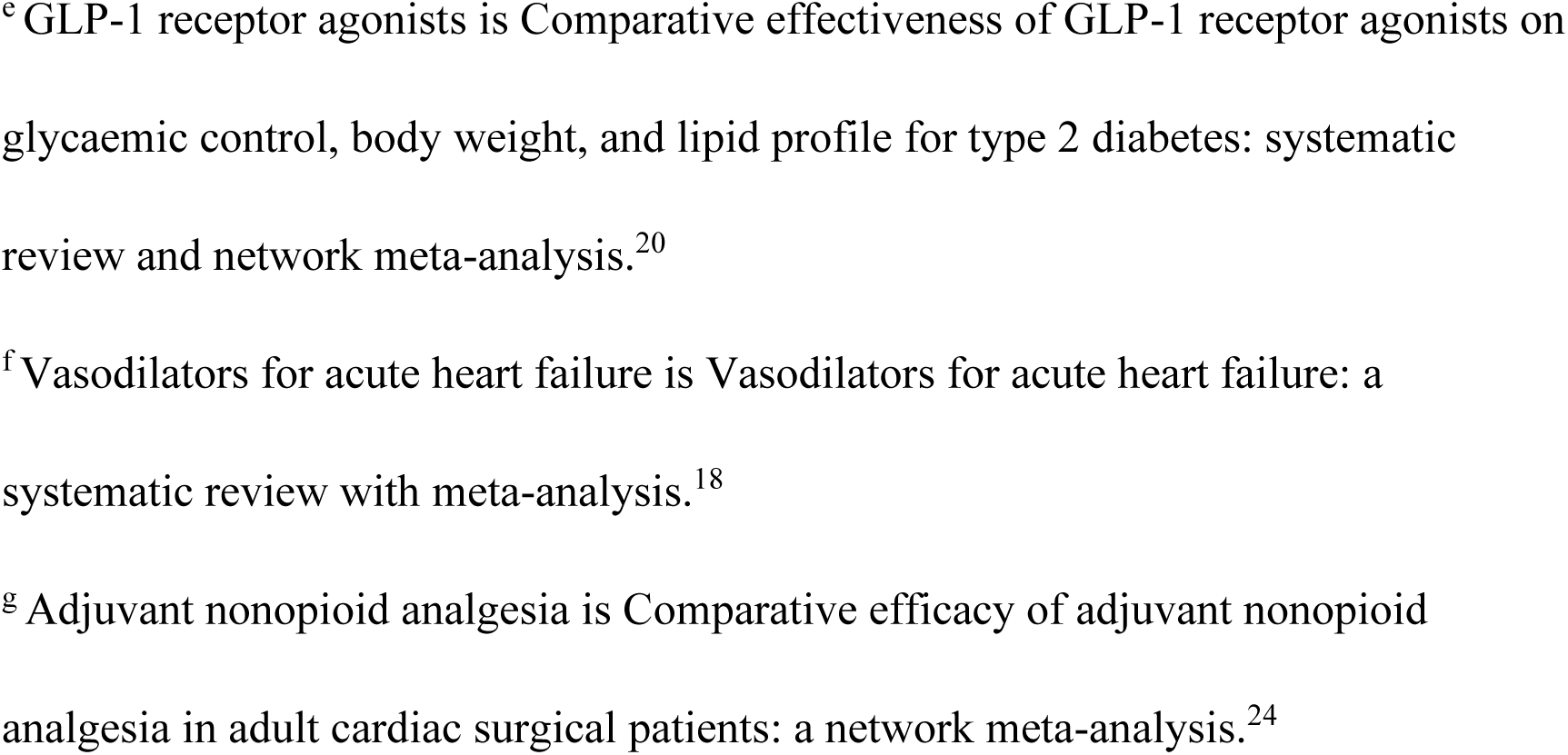
Performance of MedRAGent with DeepSeek-V3-0324 in Screening Titles and Abstracts.

| Study | n/N<sup>a</sup> | Sensitivity | Specificity | G-mean |
| --- | --- | --- | --- | --- |
| Type 1 diabetes<sup>b</sup> | 21/37 | 0.9130 | 0.9286 | 0.9208 |
| Prolonged vs. intermittent infusions<sup>c</sup> | 38/1569 | 0.8444 | 0.9875 | 0.9132 |
| Insomnia disorder<sup>d</sup> | 503/6247 | 0.8687 | 0.9795 | 0.9224 |
| GLP-1 receptor agonists<sup>e</sup> | 257/10457 | 0.7060 | 0.9896 | 0.8359 |
| Vasodilators for acute heart failure<sup>f</sup> | 82/10972 | 0.8723 | 0.9745 | 0.9220 |
| Adjuvant nonopioid analgesia<sup>g</sup> | 269/21317 | 0.8054 | 0.9991 | 0.8970 |
| Total | 1170/50599 | 0.8131 | 0.9891 | 0.8968 |

<sup>a</sup> n is number of records that were included by both MedRAGent using DeepSeek-V3-0324 and reviewers. N is number of retrieved records that have abstracts.
<sup>b</sup> Type 1 diabetes is Ramadan fasting among adolescents with type 1 diabetes: a systematic review and meta-analysis.<sup>16</sup>
<sup>c</sup> Prolonged vs. intermittent infusions is Prolonged vs. intermittent infusions of $\beta$ -Lactam antibiotics in adults with sepsis or septic Shock: a systematic review and meta-analysis.<sup>23</sup>
<sup>d</sup> Insomnia disorder is Comparative effects of pharmacological interventions for the acute and long-term management of insomnia disorder in adults: a systematic review and network meta-analysis.<sup>19</sup>
<sup>e</sup> GLP-1 receptor agonists is Comparative effectiveness of GLP-1 receptor agonists on glycaemic control, body weight, and lipid profile for type 2 diabetes: systematic review and network meta-analysis.<sup>20</sup>
<sup>f</sup> Vasodilators for acute heart failure is Vasodilators for acute heart failure: a systematic review with meta-analysis.<sup>18</sup>
<sup>g</sup> Adjuvant nonopioid analgesia is Comparative efficacy of adjuvant nonopioid analgesia in adult cardiac surgical patients: a network meta-analysis.<sup>24</sup>

**Table 3.**
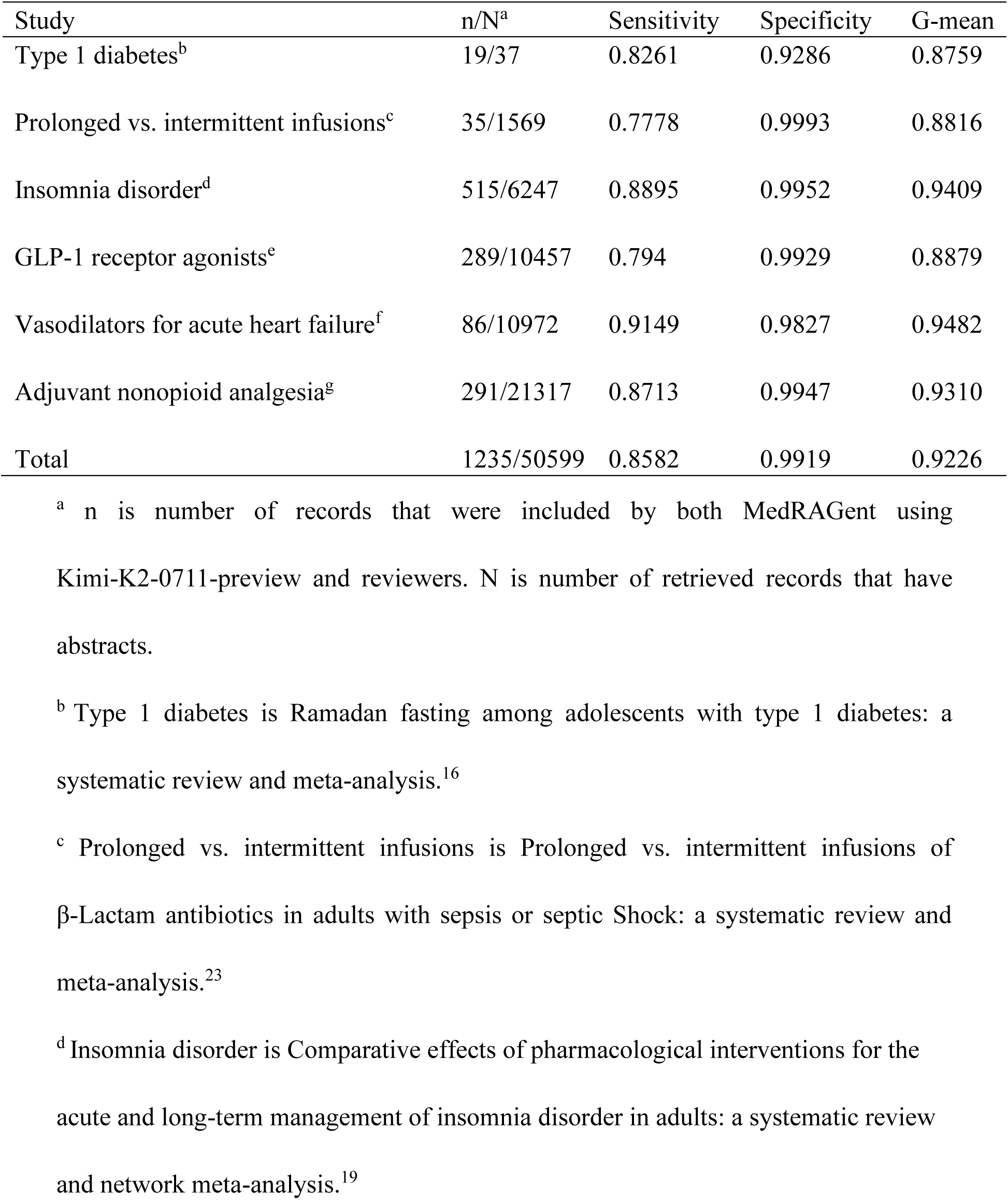
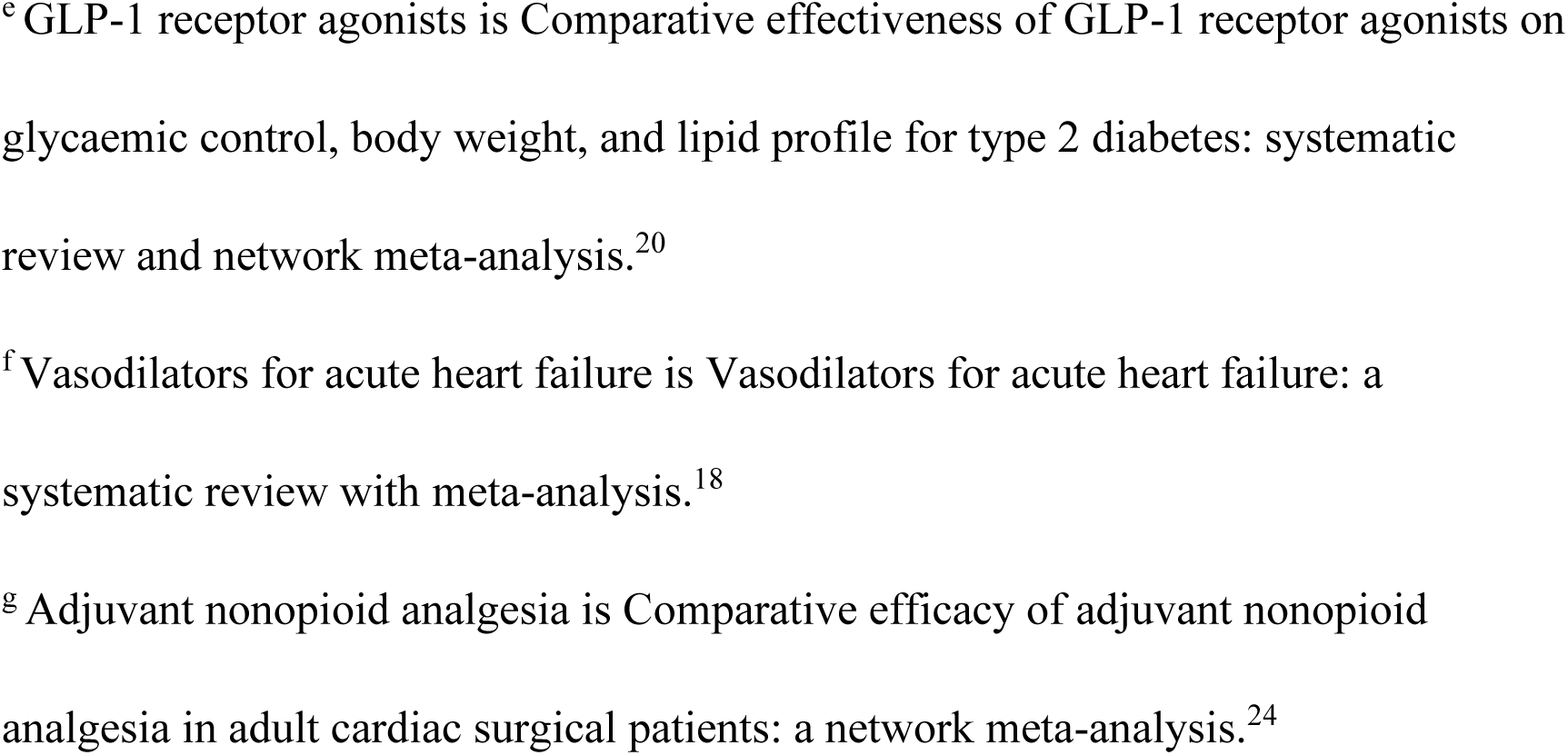
Performance of MedRAGent with Kimi-K2-0711-preview in Screening Titles and Abstracts.

The time required by MedRAGent to construct Boolean queries and conduct automatic retrieval depended on input prompts, construction rules, and retrieval scale. The retrieval time ranged from several minutes to over ten minutes. The running time and cost of MedRAGent’s automatic screening of literature titles and abstracts were related to prompt design, LLM selection, and screening volume. With DeepSeek-V3-0324, MedRAGent processed 5,000–7,000 papers daily at an average cost of 0.0025 CNY per paper. With Kimi-K2-0711-preview, it screened 4,000–5,000 papers per day at an average cost of 0.0072 CNY per paper.

## Discussion

In this study, we propose MedRAGent, a framework designed to enhance the efficiency and accuracy of systematic reviews by automating Boolean query construction and literature screening. Our results demonstrated that MedRAGent achieved competitive performance in both Boolean query construction and abstract-level screening tasks, though with some variations across different medical domains. The system exhibited its strongest precision performance (0.5227) in the Ramadan fasting study,^16^ despite showing lower recall compared to other studies. This outcome can be attributed to the highly specific free term "Ramadan" and the relatively smaller size of the dataset, which contributed to more precise retrieval while limiting comprehensive coverage. In addition, for the GLP-1 receptor agonists study,^20^ both LLMs demonstrated lower sensitivity. This performance limitation was due to the models’ difficulty in comprehending complex methodological criteria, particularly the exclusion of non-inferiority studies comparing GLP-1RA to other drugs without a placebo arm. This specific case highlights the current limitations of LLMs in interpreting complicated research design requirements. Despite these domain-specific challenges, MedRAGent maintained strong overall performance. The system achieved an overall recall of 0.8308 in Boolean query construction and specificity exceeding 0.98 in most screening tasks. The operational efficiency further underscores its practical utility, with processing capabilities of 4,000-7,000 papers daily at minimal cost.

Compared with recent work by Wang et al.,^21^ MedRAGent demonstrated superior performance in Boolean query construction. Both the DeepSeek-V3-0324 and Kimi-K2-0711-preview implementations achieved higher overall recall than the prompt-based LLM methods evaluated in their study. In comparison to the three-layer screening framework developed by Matsui et al. using GPT-4,^8^ MedRAGent showed competitive performance in abstract-level screening. While our system achieved slightly lower sensitivity in larger-scale experiments, it maintained comparable specificity levels. This performance suggests that MedRAGent offers a viable alternative for automated literature screening, particularly in scenarios requiring high specificity to minimize false inclusions.

From our results, MedRAGent with Kimi-K2-0711-preview demonstrated superior performance across multiple metrics compared to the DeepSeek-V3-0324 implementation, achieving higher values in overall sensitivity, specificity, G-mean and other metrics (Table 2 and Table 3; eTable 7 in Supplement 1). These differences in performance align with the relative rankings of these models on the Language Model Arena (LMArena), an open platform for evaluating AI systems through human preference assessment.^22^ Besides the DeepSeek-V3-0324 and Kimi-K2-0711-preview used by MedRAGent, other LLMs also have the potential to accelerate and simplify the literature screening process. This efficiency improvement can save valuable time and energy for researchers, allowing them to focus on more complex tasks, thereby promoting consistency in the selection of relevant papers and reducing the possibility of human errors and biases. This increase in consistency level can, in turn, enhance the overall quality of the evidence synthesized in clinical reviews, providing a more solid foundation for medical decision-making and the formulation of clinical guidelines.^14^

### Limitations

Although MedRAGent demonstrates potential and possibility of automated systematic review, its limitations and challenges cannot be ignored. First, we observed a performance gap between the system’s high specificity in excluding irrelevant papers and its relatively lower sensitivity in including relevant ones. This gap indicates that although the LLMs in MedRAGent can effectively exclude irrelevant papers, it lacks the understanding as proficient researchers in identifying all pertinent literatures. Our study suggests that this limitation is due to the lack of domain-specific knowledge and outdated information in the training data of the LLMs, leading to errors in the assessment of disease classifications and experimental methodologies. These limitations sometimes result in the omission of important studies, potentially affecting review comprehensiveness. Furthermore, because a small number of the exported articles through the E-utilities API lack abstracts, MedRAGent did not screen these articles. Therefore, the current LLM may not be considered as a complete substitute for human researchers in conducting systematic reviews. Instead, it functions best as a support tool that helps researchers construct Boolean queries, provides references, and expediates the literature screening process, thus allowing researchers to devote more attention to complex and judgment-intensive aspects of the review process.

## Conclusions

In conclusion, MedRAGent proves to be a valuable tool for constructing Boolean queries and performing abstract-level relevance screening for clinical review papers. By automating repetitive and time-intensive tasks, MedRAGent not only enhances efficiency but also allows researchers to concentrate on higher-level analytical responsibilities. It thus serves as a practical support tool for academic and clinical teams undertaking systematic reviews and meta-analyses, facilitating more rigorous and efficient clinical research.

## Data Availability

Code is available at GitHub (URL: https://github.com/czczy/MedRAGent). The repository will be made publicly accessible upon preprint publication. The datasets used and analyzed in this study are available from the corresponding author upon reasonable request.

