## Supplement 1 for "MedRAGent: An Automatic Literature Retrieval and Screening System Utilizing Large Language Models with Retrieval-Augmented Generation"

**Supplemental Online Content**

**eTextbox 1.** Prompt for Extracting Keywords

**eTextbox 2.** MESH Term and Supplementary Term Retrieval

**eTextbox 3.** Synonymous Terms Expansion

**eTextbox 4.** Boolean Queries Construction

**eTextbox 5.** Prompt for Literature Abstract-Level Relevance Screening

**eTable 1.** List of Population of Inclusion Criteria for Literature Screening

**eTable 2.** List of Intervention of Inclusion Criteria for Literature Screening

**eTable 3.** List of Comparison of Inclusion Criteria for Literature Screening

**eTable 4.** List of Outcome of Inclusion Criteria for Literature Screening

**eTable 5.** List of Study Design of Inclusion Criteria for Literature Screening

**eTable 6.** List of Exclusion Criteria for Literature Screening

**eTable 7.** Statistics of MedRAGent with DeepSeek-V3-0324 and Kimi-K2-0711-preview in Screening Titles and Abstracts

**eReferences.**

**eTextbox 1. Prompt for Extracting Keywords**

Please extract all nouns from the text, following these rules:

1. Keep compound nouns intact (e.g., randomised clinical trials → Randomized Controlled Trials, before surgery → before surgery)

2. Do not split phrases connected by conjunctions

3. Output one noun per line

4. No explanation needed

Text content: {text}

**eTextbox 2. MESH Term and Supplementary Term Retrieval**

=== Processing P^a^ Module ===

Extracted nouns: ['Type 1 diabetes mellitus', 'Adolescent']

[Main index match] Type 1 diabetes mellitus → Diabetes Mellitus, Type 1 (similarity: 0.7044)

[Main index match] Adolescent → Adolescent (similarity: 0.7538)

=== Processing I^b^ Module ===

Extracted nouns: ['Fasting', 'Ramadan']

[Main index match] Fasting → Fasting (similarity: 0.7068)

[No match] Ramadan Main index similarity: 0.4574, Supplementary index similarity: 0.5852

^a^ P is population.

^b^ I is intervention.

**eTextbox 3. Synonymous Terms Expansion**

=== Processing P^a^ Module ===

Term expansion: Type 1 diabetes mellitus → ['Diabetes Mellitus, Type 1', 'Autoimmune Diabetes', 'Diabetes Mellitus, Brittle', 'Diabetes Mellitus, Insulin-Dependent', 'Diabetes Mellitus, Insulin-Dependent, 1', 'Diabetes Mellitus, Juvenile-Onset', 'Diabetes Mellitus, Ketosis-Prone', 'Diabetes Mellitus, Sudden-Onset', 'Diabetes Mellitus, Type I', 'Diabetes, Autoimmune', 'IDDM', 'Insulin-Dependent Diabetes Mellitus 1', 'Juvenile-Onset Diabetes', 'Type 1 Diabetes', 'Type 1 Diabetes Mellitus']

Term expansion: Adolescent → ['Adolescent', 'Adolescence', 'Adolescents', 'Adolescents, Female', 'Adolescents, Male', 'Teenagers', 'Teens', 'Youth']

=== Processing I^b^ Module ===

Term expansion: Fasting → ['Fasting', 'Hunger Strike']

^a^ P is population.

^b^ I is intervention.

**eTextbox 4. Boolean Queries Construction**

Generated search query:

((Diabetes, Autoimmune OR Diabetes Mellitus, Type 1 OR Type 1 Diabetes Mellitus OR Diabetes Mellitus, Brittle OR IDDM OR Insulin-Dependent Diabetes Mellitus 1 OR Diabetes Mellitus, Insulin-Dependent, 1 OR Diabetes Mellitus, Sudden-Onset OR Autoimmune Diabetes OR Diabetes Mellitus, Juvenile-Onset OR Type 1 Diabetes OR Diabetes Mellitus, Ketosis-Prone OR Diabetes Mellitus, Insulin-Dependent OR Diabetes Mellitus, Type I OR Juvenile-Onset Diabetes) AND (Teens OR Youth OR Adolescence OR Adolescent OR Adolescents, Male OR Adolescents OR Adolescents, Female OR Teenagers)) AND ((Hunger Strike OR Fasting) AND (Ramadan)) AND "2001/05/01"[Date - Publication] : "2024/02/13"[Date - Publication]

**eTextbox 5. Prompt for Literature Abstract-Level Relevance Screening**

Literature Screening Rules Explanation:
1. Inclusion Criteria Handling Principles:
 - All inclusion criteria must be met → Include the Literature
 - If the abstract and title contain explicit supportive descriptions → Consider the inclusion criterion met
 - If the abstract and title contain explicit negative descriptions → Consider the inclusion criterion as unmet
 - If no explicit supportive/negative description was found in the abstract and title → The inclusion criterion is considered met

2. Exclusion Criteria Handling Principles:
 - Meets any exclusion criterion → Exclude the Literature
 - If the abstract and title contain explicit supportive descriptions → Consider the exclusion criterion met
 - If the abstract and title contain explicit negative descriptions → Consider the exclusion criterion as unmet
 - If no explicit supportive/negative descriptions are found in the abstract and title → The exclusion criteria is considered unmet

Analysis Criteria:
Inclusion Criteria:
{format_criteria(inclusion_criteria)}
Exclusion Criteria:
{format_criteria(exclusion_criteria)}

You are a senior biostatistician. Please analyze whether the following literature meets the criteria according to the steps, rules, and standards above:
1. Check each inclusion criterion one by one:
- For each inclusion criterion, search for supportive/negative descriptions in the abstract and title, then judge the literature based on the inclusion criteria handling principles.
2. Check exclusion criteria:
- For each exclusion criterion, search for supportive/negative descriptions in the abstract and title, then judge the literature based on the exclusion criteria handling principles.
3. Final judgment:
- All inclusion criteria met + no exclusion criteria met → Include (true)
- Any inclusion criterion unmet → Exclude (false)
- Any exclusion criterion met → Exclude (false)

4. Please return the result in English.

Literature Abstract and Title:
{article['Abstract'], article['Title']}

**eTable 1. List of Population of Inclusion Criteria for Literature Screening**

| **Study** | **Population** |
| --- | --- |
| Type 1 diabetes^a^ | Type 1 diabetes mellitus, adolescent. |
| Prolonged vs. intermittent infusions^b^ | Critically ill patients ($\geq$18 years old) or adult ICU patients with sepsis or septic shock or systemic inflammatory response syndrome receiving care in the intensive care unit (ICU). |
| Insomnia disorder^c^ | Adults with a diagnosis of insomnia disorder or sleep disorder or sleep initiation and maintenance disorder or sleep wake disorder or insomnia or sleep disorders. |
| GLP-1 receptor agonists^d^ | Adults with type 2 diabetes, regardless of sex, nationalities. |
| Vasodilators for acute heart failure^e^ | Adults with acute heart failure or pulmonary edema or dyspnea or decompensated congestive heart failure or decompensated heart failure or heart failure or pulmonary oedema or ventricular failure, regardless of age, sex, or comorbidities. |
| Adjuvant nonopioid analgesia^f^ | Adults (≥18 years of age) undergoing cardiac surgery (cardiac surgery includes the following, heart, mitra-valve, tricuspid-valve, pulmonary-valve, heart-lung, cardioplegia, aortic-valve-implant, heart-valve-prothesis-implant, myocardial revascularization, angioplasty, balloon, coronary, coronary-atherectom, rotational-atherectom, mammary-artery-anastomos, pericardial-window, pericardiostom, pericardiectom, pericardiocentes, tavr, aortic-root, left-venticular-assist-device, extracorporeal-membrane-oxygenation, extracorporeal membrane oxygenation, heart neoplasms, cardiac-neoplasm, atrial-myxoma, aortic-aneurysm, root-aneurysm, arch-aneurysm, thoracic-aneurysm, thoraco-abdominal-aneurysm, aortic-dissection, thoracic surgery, cardiovascular surgical procedures, cardiac surgical procedures, cardiac surgery, artery bypass surgery, cardiac surgeries, CABG, cardiopulmonary bypass, coronary artery bypass grafting, myocardial surgery, mitral valve replacement surgery, post-cardiac surgery, coronary artery bypass graft surgery, valvular heart surgery, artery by-pass surgery, heart surgery, chronic stable angina, artery bypass graft, coronary artery surgery, postcardiac surgery, surgical correction, aortic surgery, coronary surgery, post-cardiopulmonary bypass, SIRS, cardiac valve replacement, coronary artery bypass graft surgery with cardiopulmonary bypass, cardiac surgery with cardiopulmonary bypass). |

^a^ Type 1 diabetes is Ramadan fasting among adolescents with type 1 diabetes: a systematic review and meta-analysis.^1^

^b^ Prolonged vs. intermittent infusions is prolonged vs. intermittent infusions of β-Lactam antibiotics in adults with sepsis or septic shock: a systematic review and meta-analysis.^2^

^c^ Insomnia disorder is comparative effects of pharmacological interventions for the acute and long-term management of insomnia disorder in adults: a systematic review and network meta-analysis.^3^

^d^ GLP-1 receptor agonists is comparative effectiveness of GLP-1 receptor agonists on glycaemic control, body weight, and lipid profile for type 2 diabetes: systematic review and network meta-analysis.^4^

^e^ Vasodilators for acute heart failure is vasodilators for acute heart failure: a systematic review with meta-analysis.^5^

^f^ Adjuvant nonopioid analgesia is comparative efficacy of adjuvant nonopioid analgesia in adult cardiac surgical patients: a network meta-analysis.^6^

**eTable 2. List of Intervention of Inclusion Criteria for Literature Screening**

| **Study** | **Intervention** |
| --- | --- |
| Type 1 diabetes^a^ | Fasting during Ramadan. |
| Prolonged vs. intermittent infusions^b^ | Prolonged infusion of a beta-lactam antibiotic (the beta-lactam antibiotic includes the following, β-lactam antibiotic, meropenem, carbapenem, cephalosporin, monobactam, penicillin), where “prolonged infusion” is defined as either:  Extended infusion: intravenous drug administration for $\geq$2 hours during a dosing interval.  Continuous infusion or continuous administration or continuous application: constant intravenous drug administration either as a sequential 6-hour, 8-hour, 12-hour or 24-hour infusion. |
| Insomnia disorder^c^ | Pharmacological treatments as combination therapy or monotherapy (pharmacological treatments including alprazolam, amitriptyline, antihistamine, anti-histamine, doxepin, doxylamine, diphenhydramine, mirtazapine, trazodone, benzodiazepine, brotizolam, clonazepam, diazepam , estazolam, lunitrazepam, flurazepam, haloxazolam, hydroxyzine, loprazolam, lorazepam, lormetazepam, midazolam, nimetazepam, nitrazepam, oxazepam, quazepam, quetiapine, promethazine, rilmazafone, temazepam, triazolam, nonbenzodiazepine, non-benzodiazepine, Z drug, Z drugs, eszopiclone, zaleplon, zolpidem, zopiclone, melatonine, ramelteon, orexin, suvorexant, lemborexant, hypnotics, daridorexant, polysomnography, gaboxadol, trimipramine). |
| GLP-1 receptor agonists^d^ | GLP-1 receptor agonists (monotherapy or added to non-randomised background hypoglycaemic treatments, hypoglycaemic treatments include the following, insulin glargine, insulin degludec, cagrilintide, sitagliptin. The GLP-1 receptor agonists include the following, liraglutide, semaglutide, exenatide, dulaglutide, loxenatide, efpeglenatide, lixisenatide, orforglipron, tirzepatide, mazdutide, albiglutide, retatrutide, iGlarLixi, ITCA 650, LY3437943, PEX168). |
| Vasodilators for acute heart failure^e^ | Vasodilator drugs, vasodilator agents, antihypertensive agents, calcium channel blockers, nitric oxide donors, nitroglycerin, isosorbide dinitrate, relaxin, nesiritide, nitroglycerin, serelaxin , isosorbide dinitrate, vasodilator therapy, captopril, prazosin, tezosentan, enalaprilat, clevidipine, ularitide, urapidil. |
| Adjuvant nonopioid analgesia^f^ | Nonopioid analgesics (Nonopioid analgesics includes the following, e.g., analgesia, analgesics, corticosteroids, dexmedetomidine, dexamethasone, acetaminophen, ketamine, magnesium, methylprednisolone, nonsteroidal anti-inflammatory medications, diclofenac, clonidine, corticosteroids, aspirin, hydrocortisone, glucocorticosteroids, steroids, magnesium sulfate, pregabalin, cyclooxygenase 2 Inhibitors, tirilazad mesylate, atorvastatin. |

^a^ Type 1 diabetes is Ramadan fasting among adolescents with type 1 diabetes: a systematic review and meta-analysis.^1^

^b^ Prolonged vs. intermittent infusions is prolonged vs. intermittent infusions of β-Lactam antibiotics in adults with sepsis or septic shock: a systematic review and meta-analysis.^2^

^c^ Insomnia disorder is comparative effects of pharmacological interventions for the acute and long-term management of insomnia disorder in adults: a systematic review and network meta-analysis.^3^

^d^ GLP-1 receptor agonists is comparative effectiveness of GLP-1 receptor agonists on glycaemic control, body weight, and lipid profile for type 2 diabetes: systematic review and network meta-analysis.^4^

^e^ Vasodilators for acute heart failure is vasodilators for acute heart failure: a systematic review with meta-analysis.^5^

^f^ Adjuvant nonopioid analgesia is comparative efficacy of adjuvant nonopioid analgesia in adult cardiac surgical patients: a network meta-analysis.^6^

**eTable 3. List of Comparison of Inclusion Criteria for Literature Screening**

| **Study** | **Comparison** |
| --- | --- |
| Type 1 diabetes^a^ | NA^g^ |
| Prolonged vs. intermittent infusions^b^ | Intermittent infusion of a beta-lactam antibiotic where “intermittent infusion” is defined as administration of an intravenous drug infusion for <2 hours. |
| Insomnia disorder^c^ | Placebo or another active pharmacological agent or psychotherapy. |
| GLP-1 receptor agonists^d^ | Placebo or any GLP-1 receptor agonists. |
| Vasodilators for acute heart failure^e^ | Placebo, no intervention, or another active treatment. |
| Adjuvant nonopioid analgesia^f^ | Other active nonopioid analgesics, placebo, or no additional treatment. |

^a^ Type 1 diabetes is Ramadan fasting among adolescents with type 1 diabetes: a systematic review and meta-analysis.^1^

^b^ Prolonged vs. intermittent infusions is prolonged vs. intermittent infusions of β-Lactam antibiotics in adults with sepsis or septic shock: a systematic review and meta-analysis.^2^

^c^ Insomnia disorder is comparative effects of pharmacological interventions for the acute and long-term management of insomnia disorder in adults: a systematic review and network meta-analysis.^3^

^d^ GLP-1 receptor agonists is comparative effectiveness of GLP-1 receptor agonists on glycaemic control, body weight, and lipid profile for type 2 diabetes: systematic review and network meta-analysis.^4^

^e^ Vasodilators for acute heart failure is vasodilators for acute heart failure: a systematic review with meta-analysis.^5^

^f^ Adjuvant nonopioid analgesia is comparative efficacy of adjuvant nonopioid analgesia in adult cardiac surgical patients: a network meta-analysis.^6^

^g^ NA represents null values.

**eTable 4. List of Outcome of Inclusion Criteria for Literature Screening**

| **Study** | **Outcome** |
| --- | --- |
| Type 1 diabetes^a^ | Hyperglycemia, hypoglycemia, diabetic ketoacidosis, changes in HbA1c, and weight changes |
| Prolonged vs. intermittent infusions^b^ | All-cause 90-day mortality, if 90-day mortality outcomes are not reported in a study, we will use the time closest to day 90 (before and beyond). ICU mortality, ICU length of stay as reported in the original study, clinical cure as defined in the original study, microbiological cure as defined in the original study, adverse events as defined in the original publication |
| Insomnia disorder^c^ | Patient-rated quality of sleep, all-cause discontinuation, discontinuation due to adverse events, number of patients with $\geq$1 adverse event, sleep onset latency, wake time after sleep onset, total sleep time, number of awakenings, hangover, rebound/withdrawal phenomena, specific adverse events. |
| GLP-1 receptor agonists^d^ | Changes from baseline in outcomes of HbA1c concentrations, fasting blood glucose concentrations, body weight, body mass index, waist circumference, and serum lipid parameters (e.g., high density lipoprotein, low density lipoprotein, total cholesterol, and triglyceride levels); adverse events (e.g., treatment discontinuation due to adverse events, all-cause death, cardiovascular disease, non-fatal stroke, kidney failure, severe hypoglycaemia, eye disease requiring intervention, health-related quality of life, serious gastrointestinal events, etc.). |
| Vasodilators for acute heart failure^e^ | All-cause mortality; serious adverse events (SAEs), tracheal intubation, length of hospital-stay, blood pressure reduction. |
| Adjuvant nonopioid analgesia^f^ | Resting postoperative pain scores, postoperative opioid consumption in morphine equivalents (total postoperative morphine dose), lengths of intensive care unit (ICU), lengths of hospital stay, duration of mechanical ventilation; incidences of myocardial infarction, delirium, nausea, vomiting, pharmacological and mechanical support, hypotension, hypertension, hyperdynamic responses, muscle rigidity, postoperative shivering, tachycardia, additional doses, increment, adverse events, extubation time, Atrial fibrillation, kidney injury, arrhythmia, Inflammatory markers (IL-6, PNE). |

^a^ Type 1 diabetes is Ramadan fasting among adolescents with type 1 diabetes: a systematic review and meta-analysis.^1^

^b^ Prolonged vs. intermittent infusions is prolonged vs. intermittent infusions of β-Lactam antibiotics in adults with sepsis or septic shock: a systematic review and meta-analysis.^2^

^c^ Insomnia disorder is comparative effects of pharmacological interventions for the acute and long-term management of insomnia disorder in adults: a systematic review and network meta-analysis.^3^

^d^ GLP-1 receptor agonists is comparative effectiveness of GLP-1 receptor agonists on glycaemic control, body weight, and lipid profile for type 2 diabetes: systematic review and network meta-analysis.^4^

^e^ Vasodilators for acute heart failure is vasodilators for acute heart failure: a systematic review with meta-analysis.^5^

^f^ Adjuvant nonopioid analgesia is comparative efficacy of adjuvant nonopioid analgesia in adult cardiac surgical patients: a network meta-analysis.^6^

**eTable 5. List of Study Design of Inclusion Criteria for Literature Screening**

| **Study** | **Study design** |
| --- | --- |
| Type 1 diabetes^a^ | Observational studies (e.g., prospective study, cohort studies, case-control studies, cross-sectional studies). |
| Prolonged vs. intermittent infusions^b^ | Randomized controlled trials, pilot study (Randomized controlled trials include the following, randomized controlled trial, controlled clinical trial, random, placebo, randomized, controlled trial, double-blind, single-blind). |
| Insomnia disorder^c^ | Randomized controlled trials, pilot study (Randomized controlled trials include the following, randomized controlled trial, controlled clinical trial, random, placebo, randomized, controlled trial, double-blind). |
| GLP-1 receptor agonists^d^ | Randomized controlled trials, pilot study (Randomized controlled trials include the following, randomized controlled trial, controlled clinical trial, random, placebo, randomized, controlled trial, double-blind, single-blind). |
| Vasodilators for acute heart failure^e^ | Randomized controlled trials, retracted publication, retraction of publication, pilot study (randomized controlled trials include the following, randomized controlled trial, controlled clinical trial, random, placebo, randomized, controlled trial, double-blind, single-blind). |
| Adjuvant nonopioid analgesia^f^ | Randomized controlled trials, pilot study (randomized controlled trials include the following, randomized controlled trial, controlled clinical trial, random, placebo, randomized, controlled trial, double-blind, single-blind). |

^a^ Type 1 diabetes is Ramadan fasting among adolescents with type 1 diabetes: a systematic review and meta-analysis.^1^

^b^ Prolonged vs. intermittent infusions is prolonged vs. intermittent infusions of β-Lactam antibiotics in adults with sepsis or septic shock: a systematic review and meta-analysis.^2^

^c^ Insomnia disorder is comparative effects of pharmacological interventions for the acute and long-term management of insomnia disorder in adults: a systematic review and network meta-analysis.^3^

^d^ GLP-1 receptor agonists is comparative effectiveness of GLP-1 receptor agonists on glycaemic control, body weight, and lipid profile for type 2 diabetes: systematic review and network meta-analysis.^4^

^e^ Vasodilators for acute heart failure is vasodilators for acute heart failure: a systematic review with meta-analysis.^5^

^f^ Adjuvant nonopioid analgesia is comparative efficacy of adjuvant nonopioid analgesia in adult cardiac surgical patients: a network meta-analysis.^6^

**eTable 6. List of Exclusion Criteria for Literature Screening**

| **Study** | **Exclusion criteria** |
| --- | --- |
| Type 1 diabetes^a^ | Animal studies. Case reports. Systematic review. Meta-analysis. |
| Prolonged vs. intermittent infusions^b^ | Retrospective cohort studies. Meta-analysis. Systematic review. |
| Insomnia disorder^c^ | Meta-analysis. Systematic review. Cluster-randomized trials. Crossover trials. |
| GLP-1 receptor agonists^d^ | Crossover design trials. Conference abstracts. Meta-analysis. Systematic review.  Non-inferiority studies comparing GLP-1RA to other drug classes without a placebo arm.  Using withdrawn drugs. |
| Vasodilators for acute heart failure^e^ | Trials using inodilators as intervention (e.g., levosimendan, dobutamine, milrinone).  Trials that included patients with cardiogenic shock.  Meta-analysis. Systematic review. |
| Adjuvant nonopioid analgesia^f^ | Trials studying nerve blocks and local anesthetics.  Trials enrolling patients undergoing minimally invasive surgery.  Meta-analysis. Systematic review. |

^a^ Type 1 diabetes is Ramadan fasting among adolescents with type 1 diabetes: a systematic review and meta-analysis.^1^

^b^ Prolonged vs. intermittent infusions is prolonged vs. intermittent infusions of β-Lactam antibiotics in adults with sepsis or septic shock: a systematic review and meta-analysis.^2^

^c^ Insomnia disorder is comparative effects of pharmacological interventions for the acute and long-term management of insomnia disorder in adults: a systematic review and network meta-analysis.^3^

^d^ GLP-1 receptor agonists is comparative effectiveness of GLP-1 receptor agonists on glycaemic control, body weight, and lipid profile for type 2 diabetes: systematic review and network meta-analysis.^4^

^e^ Vasodilators for acute heart failure is vasodilators for acute heart failure: a systematic review with meta-analysis.^5^

^f^ Adjuvant nonopioid analgesia is comparative efficacy of adjuvant nonopioid analgesia in adult cardiac surgical patients: a network meta-analysis.^6^

**eTable 7. Statistics of MedRAGent with DeepSeek-V3-0324 and Kimi-K2-0711-preview in Screening Titles and Abstracts**

|  | **TP^a^** | **FP^b^** | **FN^c^** | **TN^d^** |
| --- | --- | --- | --- | --- |
| **DeepSeek-V3-0324** | | | | |
| **Type 1 diabetes^e^** | 21 | 1 | 2 | 13 |
| **Prolonged vs. intermittent infusions^f^** | 38 | 19 | 7 | 1505 |
| **Insomnia disorder^g^** | 503 | 116 | 76 | 5552 |
| **GLP-1 receptor agonists^h^** | 257 | 105 | 107 | 9988 |
| **Vasodilators for acute heart failure^i^** | 82 | 277 | 12 | 10601 |
| **Adjuvant nonopioid analgesia^j^** | 269 | 19 | 65 | 20964 |
| **Total** | 1170 | 537 | 269 | 48623 |
| **Kimi-K2-0711-preview** | | | | |
| **Type 1 diabetes^e^** | 19 | 1 | 4 | 13 |
| **Prolonged vs. intermittent infusions^f^** | 35 | 1 | 10 | 1523 |
| **Insomnia disorder^g^** | 515 | 27 | 64 | 5641 |
| **GLP-1 receptor agonists^h^** | 289 | 72 | 75 | 10021 |
| **Vasodilators for acute heart failure^i^** | 86 | 188 | 8 | 10690 |
| **Adjuvant nonopioid analgesia^j^** | 291 | 111 | 43 | 20872 |
| **Total** | 1235 | 400 | 204 | 48760 |

^a^ TP is true positive.

^b^ FP is false positive.

^c^ FN is false negative.

^d^ TN is true negative.

^e^ Type 1 diabetes is Ramadan fasting among adolescents with type 1 diabetes: a systematic review and meta-analysis.^1^

^f^ Prolonged vs. intermittent infusions is prolonged vs. intermittent infusions of β-Lactam antibiotics in adults with sepsis or septic shock: a systematic review and meta-analysis.^2^

^g^ Insomnia disorder is comparative effects of pharmacological interventions for the acute and long-term management of insomnia disorder in adults: a systematic review and network meta-analysis.^3^

^h^ GLP-1 receptor agonists is comparative effectiveness of GLP-1 receptor agonists on glycaemic control, body weight, and lipid profile for type 2 diabetes: systematic review and network meta-analysis.^4^

^i^ Vasodilators for acute heart failure is vasodilators for acute heart failure: a systematic review with meta-analysis.^5^

^j^ Adjuvant nonopioid analgesia is comparative efficacy of adjuvant nonopioid analgesia in adult cardiac surgical patients: a network meta-analysis.^6^
